# Opinions of Former Jail Residents about Self-collection of SARS-CoV-2 Specimens, Paired with Wastewater Surveillance: A Qualitative Study Rapidly Examining Acceptability of COVID-19 Mitigation Measures

**DOI:** 10.1101/2022.11.29.22282848

**Authors:** Myrna del Mar González-Montalvo, Peter F. Dickson, Lindsay B. Saber, Rachel A. Boehm, Victoria L. Phillips, Matthew J. Akiyama, Anne C. Spaulding

**Affiliations:** Department of Global Health, Rollins School of Public Health, Emory University, Atlanta, Georgia, US; Department of Behavioral Health, Rollins School of Public Health, Emory University, Atlanta, Georgia, US; Department of Health Policy and Management, Rollins School of Public Health, Emory University, Atlanta, Georgia, US; Department of Medicine, Montefiore Hospital, New York City, New York, US; Department of Medicine, Emory School of Medicine, Atlanta, Georgia, US

**Keywords:** wastewater, surveillance, jail, covid-19, self-collection, sars-cov-2

## Abstract

In year one of the COVID-19 epidemic, the incidence of infection for US carceral populations was 5.5-fold higher than that in the community. Prior to the rapid roll out of a comprehensive jail surveillance program of Wastewater-Based Surveillance (WBS) and individual testing for SARS-CoV-2, we sought the perspectives of formerly incarcerated individuals regarding mitigation strategies against COVID-19 to inform acceptability of the new program. In focus groups, participants discussed barriers to their receiving COVID-19 testing and vaccination. We introduced WBS and individual nasal self-testing, then queried if wastewater testing to improve surveillance of emerging outbreaks before case numbers surged, and specimen self-collection, would be valued. The participants’ input gives insight into ways to improve the delivery of COVID-19 interventions. Hearing the voices of those with lived experiences of incarceration is critical to understanding their views on infection control strategies and supports including justice-involved individuals in decision-making processes regarding jail-based interventions.

## Introduction

Strict confinement in high density congregate settings increases the risk of transmission of airborne pathogens. In 2020, the incidence of infection with the severe acute respiratory syndrome coronavirus 2 (SARS-CoV-2) in US custody populations was 5.5 times higher than that in the community.(1, 2) Not only has the ability to distance been limited for persons in jails and prisons during the COVID-19 pandemic,(3, 4) but also the supply of personal protective equipment has been frequently inadequate.(5) Surveillance and good infection control practices, such as screening, confirmatory testing, isolation and quarantine, can minimize the impact of COVID-19 on both those who live and work in correctional facilities.(6)

In the 1940s, epidemiologists in the United States used wastewater-based surveillance (WBS) to detect and manage polio outbreaks.(7) The gold standard for wastewater pathogen detection, polymerase chain reaction (PCR), came on line in the 1990s and its use has continued.(8-10) Due to fatigue associated with the repeated rounds of individual swab testing several months into the COVID-19 pandemic, wastewater testing represented a way to improve surveillance of emerging outbreaks before case numbers surged. Innovative PCR-based WBS strategies have been recently emerging to monitor for SARS-CoV-2, both in private and public settings, such as university campuses,(11) neighborhoods, and now, correctional facilities.

WBS will pinpoint where outbreaks are occurring and could prompt individual testing or other mitigation measures. To decrease discomfort with individual testing, we made a preliminary decision to employ self-collected nasal swabs for the molecular testing for SARS-CoV-2 in a project planned for a local jail. The collection kit for molecular diagnostic testing for viruses was manufactured by SteriPack USA [Lakeland FL, Steripackgroup.com]. These swabs had been used for a validated molecular test with Emergency Use Authorization. Associated laboratory costs in a public health laboratory were 10%-25% of the cost of commercial laboratory testing at the time. Quality control for the method could be monitored for adequacy based on the presence of nasal epithelial cells in self-collected specimens, to ensure that those tested were cooperating with the collection procedure.

In 2006, an Institute of Medicine committee published a report on “Ethical Considerations for Research Involving Prisoners.(12, 13) It added collaboration of relevant stakeholders to the bioethical principles of justice, beneficence and respect for persons as foundational in studies involving persons in the criminal legal system. In matters of correctional health, the person with lived experience of incarceration represents the stakeholder of central importance. The goal of our present study was to understand perspectives of formerly incarcerated individuals regarding COVID-19 control strategies of jails prior to the rapid embarkment on a comprehensive surveillance and mitigation program in an Atlanta, Georgia, United States jail. Conducting focus groups to hear the voices of those with lived experiences of being held in jails and prisons during the COVID-19 pandemic is critical to understanding their views on infection control strategies.(14) Feedback from participants may inform the design of new interventions to improve outcomes among this vulnerable population.

## Methods

We employed rapid qualitative analysis, which provides timely results to tailor interventions to the needs of a target population, support iterative program improvements and provide information to health care stakeholders on a short timeline.(15-18) Authors (MDG, PFD, LBR, VLP, MJA, ACS) contributed to the development of a script for the focus group sessions. The script focused on participant experiences with mitigation measures such as masking, quarantine, isolation, vaccination, and testing in correctional settings during COVID-19. Participants were instructed in the use of self-collection of nasal specimens and swabbed themselves. They were also shown a video explaining water-based surveillance. Questions about the acceptability and the value of pairing individual testing with SARS-CoV-2 WBS as an integrated strategy followed.

Recruitment of a convenience sample of adults with a history of detention or imprisonment began in September of 2021. We approached staff of a community center established by the Atlanta Police Foundation in the neighborhood of Fulton County Jail, in northwest Atlanta, to refer persons in the neighborhood who had been in any US jail or prison at any point between March 2020 and September 2021. The center’s staff invited interested individuals who had lived experience of being incarcerated to attend one of three focus groups.

The groups convened in a conference room at the community center. Participants sat around a table while practicing social distancing and wearing masks. The groups were facilitated by Emory staff who obtained formal written consent to participate and have the session recorded. Collection of demographics occurred before the start of each focus group; participants could opt out of reporting specific personal information. Participants were compensated with a $50 gift card at the close of each focus group or when a subject asked to leave. Facilitators followed the script to elicit perspectives of the participants towards various mitigation measures. The recording of the session was professionally transcribed.

As the focus group began, each participant received one SteriPack swab. The kit consists of a 50mm polypropylene stick and a 100% Polyester bud, as well as a cylinder-like receptacle container. After a demonstration of using the swabs for specimen self-collection, the participants tried swabbing their anterior nares with the devices. We then solicited participants’ attitudes on its usability and practicality when coupled with WBS during incarceration.

To learn about WBS, participants listened to a short video on WBS on college campuses. At the time of the study, we were unable to locate previously produced videos on WBS in correctional settings. We solicited participants’ opinions on whether they thought such technology was appropriate for correctional facilities and would protect incarcerated residents.

After the transcriptions from the three focus groups were completed, we used the Rapid Assessment Process—an intensive, team-based combination inductive/deductive approach for qualitative data analysis that employs triangulation and iterative data analysis, and permits more timely delivery of findings to the field than traditional analysis.(19) Two independent coders (MGM and PFD) reviewed and summarized transcripts using a structured analysis template in Microsoft Word. Templates were organized around themes selected *a priori* from the interview guide and revised to include emergent themes as analysis unfolded. Templates, including extracted quotes, were then iteratively reviewed by the qualitative study lead (MJA) and the rest of the team to consolidate and distinguish themes. This process ensured alignment and resolved discrepancies. Final templates were used to generate and refine key learnings and identify themes with core qualitative investigators (MGM, PFD, MJA). In the key quotes presented, we italicize text from the interviewer for clarity.

The Emory University IRB approved the study protocol.

## Results

We conducted three focus groups with three, ten, and seven participants, respectively, with 17 participants identifying as men and 3 as women. Ages ranged from 18 to 67 years. All participants identified as non-Hispanic, and Black or African-American. Educational level attained ranged from tenth grade to two years of college. Two had been in prison, while the remainder reported time only in jail. Subjects were held in correctional facilities of the City of Atlanta, three different Georgia counties, one county in another state, Georgia Department of Corrections and the Federal Bureau of Prisons. Releases were between March 2020 and August 2021. The longest time spent in custody within the pandemic period was four months.

### Attitudes toward and experiences with testing

Attitudes regarding testing were positive overall, as many felt this was a good way to maintain control and prevent COVID-19 from spreading to other residents within the correctional facility. On many occasions, residents “guessed” who had tested positive because an individual was moved from their regular unit to another.

> They said we had COVID, but once they quarantined [you] in isolation by [your] self, [then] they came in and tested, [if they tested and] you had COVID [they] tested again until we better. I guess they said [they tested] until [they were negative and] they sent [us] back to the general population.

In one correctional system, former residents complained of inability to access a test unless symptomatic:

> Because if you’re not like literally like on the verge of dying or if it’s not [serious]…

There were, however, comments from participants suggesting mistrust regarding COVID-19 testing and this being associated with their race.

> But I feel like since y’all are making easier ways, that’s going to bring you better progress…. Black people, we’re not going to do it.

Another participant expressed that many were suspicious about testing due to concerns about swabs being used to implant microchips.

> A lot of people be thinking that they – they’re going some type – some type of chip in it.

Some participants mentioned hesitancy to be tested because of the stigma associated with testing positive for COVID-19. However, other participants answered that the possibility of receiving a positive COVID-19 result would not prevent them from taking the test because they ultimately would like to know if they had COVID-19 and would not want to spread the infection to those with comorbidities.

> I wouldn’t avoid. I would take it, -- You know, because you know, that’s life or death. You know, I wouldn’t want to get around no one. I would want them to, you know, put me in, you know, separate room from someone until I, you know, can get better.
>
> Because other people got asthma, you know, heart problems…different kind of…health problems that they, you know, might harm them or so yeah, I would try to have them removed – have me separate.

### Experiences with Mitigation: Masks, Quarantine and Isolation

Some participants shared that if anyone tested positive, they were isolated along with others who had also tested positive. Once a negative result came back after one or two weeks, they were sent back to the general population. Regarding experiences with masking, some participants reported significant distress before distribution of masks was widespread. One participant mentioned that correctional officers were prioritized to receive masks before residents:

> At first everybody, they was panicking. They didn’t know what to do. You know, eventually they started getting it from the higher uppers, and then they started like giving us masks. The officers had masks before we did.

Other participants received masks, either when being apprehended or after they arrived at jail intake. However, one participant noted they received a mask once and no more were received during the participant’s stay at the correctional facility:

> They gave me a mask as they were arresting me. I was in handcuffs… *And they didn’t give you another mask after that?*… No.

Reported experiences suggested some facilities placed people in quarantine for 2 weeks after intake. Other jails did not sustain entry cohort quarantining for the full SARS-CoV-2 incubation period recommended at the time. Instead, participants were separated for the first 3 days with others that entered the facility at the same time. After 3 days, they were issued jail uniforms and sent to general population.

> As soon as I went in the door it was the early state of the pandemic. They put me on lockdown…. Like they put me in lockdown, so I didn’t have to be around nobody, no nothing. No nothing. Because it was coronavirus –

In some jails infected persons were not removed from the exposed. Participants reported that ‘quarantine’ in these facilities consisted of keeping the residents of a housing unit together as one cohort, with a mix of infected and exposed individuals, to which no others were added. Only if a person was unstable were they removed:

> … So once he finally like fell out, that’s when they had let him out of the [block] – somebody had came and got him.

Some participants stated they felt the care they received while in isolation and quarantine was inadequate. Quarantine resembled punitive solitary confinement, without an effort to cohort individuals by infection status or similar exposure.

> [They] just kept me in my room…some food. I was dying of thirst [during] lockdown. They didn’t even put me around people.

Other participants mentioned it felt like a normal housing assignment to be in a quarantined cell block, just as if they were moved from one general housing unit to another.

> I mean, like where I was quarantined at it didn’t feel like a punishment. It just felt like, alright, now I’m in a new pod, and I’ve just got to meet new people now.

### Experiences with COVID-19 vaccine

Attitudes towards vaccines were divided; some opted out of vaccination when offered and others were not in jail when vaccines became available but decided to pursue vaccination in the outside community.

> You got to have it. I’m telling you. I watch the news.

> Y’all, I have took the vaccine. Personally, I did not care for it, but I took it [outside]… Because … health is my wealth, personally.

Some participants reported hesitancy toward the COVID-19 vaccine. Reasons for vaccine hesitancy included mistrust in the speed of vaccine production, concern that vaccine-related protection was not adequate, and belief in divine protection:

> How they come up with a vaccine in one year – How they going to come up with a vaccine in one year and now in one month they come up with something 12 under?
>
> They did at one point offer us vaccines. I specifically didn’t take it because I’ve got my own views on it. I wouldn’t see why – I’m trying to avoid this. Why would I put it in my body? And then on top of that, people still die with it, and just having the vaccine only lessens the symptoms.
>
> I ain’t getting the vaccine… you feel me? I don’t put nobody above but the man up above because I’m protected by Him.

### Attitudes toward nasal self-collection for COVID-19 testing

Attitudes toward nasal self-collection were generally positive. Reasons participants thought self-collection was advantageous varied, but most centered around allowing a person to test themselves, which may for some be empowering.

> I mean, I think it’ll definitely help… There’ll be obvious draw backs, but like still at the same time I feel like if you put it in somebody’s hands it’ll make them make that decision for themselves. Like it’ll make the people who are responsible more comfortable doing it.

One participant also noted that system-wide nasal self-collection represented an opportunity for correctional officers and incarcerated persons to take action that would protect each other’s safety.

> The common denominator with both detainees and jail staff is both are in the facility for a period of time, and you know, the health hazards of catching COVID. So, you know, I mean, possibly death or a hospitalization or whatever…coming together collectively.

Nasal self-collection was also viewed to be acceptable due to the shallower depth the swab needs to be inserted into the nostril.

> Yeah, because you know how far to go up in your own nose compared to somebody else. I think that would be better.

> The other thing, it’s more like it’s going up your brain. Like it’s like in your eyeballs or something.

Additional participants stated there could be harms associated with the particular nasal self-collection kit proposed for COVID-19 testing. They thought the device could not be weaponized, but, there could be a possibility of storing small amounts of substances (drugs) or small objects in the device.

> *Looking at the device, do you think someone could do harm with it, turn it into a weapon, store something in it?*
>
> … Store something in it, yeah.

While attitudes toward nasal self-collection were mixed, some expressed that additional education and incentives would increase participation.

> I would do it say if you – for the month, if you do it for the four weeks, at the fourth week we’ll give a store bag, a $5 store bag. So now you got people to do it for four weeks.

### Acceptability of wastewater testing

Participants were shown a video on WBS prior to discussion about the method. Initial participant responses indicated a lack of understanding of WBS and study staff spent considerable time with verbal explanations regarding the concept of using wastewater to test for COVID-19. Specifically, the participants had difficulty understanding what constituted wastewater collection.

Group members expressed concern about wastewater generally. In Atlanta’s local jails and elsewhere, the sink and toilet are joined in a single unit. The participants who had been in the correctional facilities articulated that both use the same source of water, meaning the wastewater, especially since the water reportedly had an unpleasant taste.

After the facilitator addressed knowledge gaps and the participants had a better understanding of wastewater surveillance, many had expressed a positive attitude toward it. Participants thought wastewater testing was an attractive surveillance method for COVID-19. Wastewater was generally endorsed, especially the combination of wastewater and self-testing. Wastewater testing would be beneficial, easy, and feasible.

> *So then do you think that will work since it will help them maybe target their testing to one specific area? …* 1,000%.
>
> Because they can pinpoint whose pod is this coming from. It’ll be easier, and then they can just give the test, and then it would – they get taken for quarantine and try to treat them.
>
> It would be an easy process. It could help more people.
>
> It’s like definitely less time consuming, like for sure. And then it’s just one surefire way of another like, okay, every two weeks – if we do this every two weeks like this is the results we’re getting. There’s no like other barriers or like – you get what I’m saying? Everybody has the bowel movements, so you know, -- Yeah.

## Discussion

Our study with individuals who had recently been incarcerated during the COVID-19 pandemic revealed important themes on mitigation measures and COVID-19 testing and surveillance. Participants had been incarcerated in multiple facilities and in discussing protocols for masking, isolation, and quarantine, many participants reported deviations from CDC recommendations, such as quarantining for periods shorter than specified in the CDC guidance for the time period and difficulties accessing tests if asymptomatic. Absence of interventions for COVID-19 management early in the epidemic that communicated that their health was a priority seemed to be at the heart of many themes that emerged. Participants also reported a lack of confidence regarding vaccine development and its purpose, leading to vaccine hesitancy, which has been reported in other studies.(20)

There was an overall acceptance of nasal self-collection as a strategy to control COVID-19 within correctional facilities. Overall, the specimen collection using the SteriPack kit was perceived to be convenient and acceptable. Individual autonomy, convenience and confidentiality are all advantages of self-collection of samples for diagnostic testing,(21) as has been previously demonstrated in jail-based sexually transmitted infection management programs.(22) Indeed, empowerment of the person through self-testing emerged as a theme in our study.

Surveillance via measuring virus in wastewater was a challenging concept for participants to understand, given they lacked background in this area and the video introduction was short. The term “wastewater” generated confusion. The use of a joint sink-and-toilet configuration in a number of jails **(Figure 1)** may explain why some participants indicated that they believed “wastewater” was what flowed from the faucet of the combination fixture. The focus group leader clarified after the video that wastewater was sewer output and reiterated its meaning in subsequent groups.

**Figure 1.**
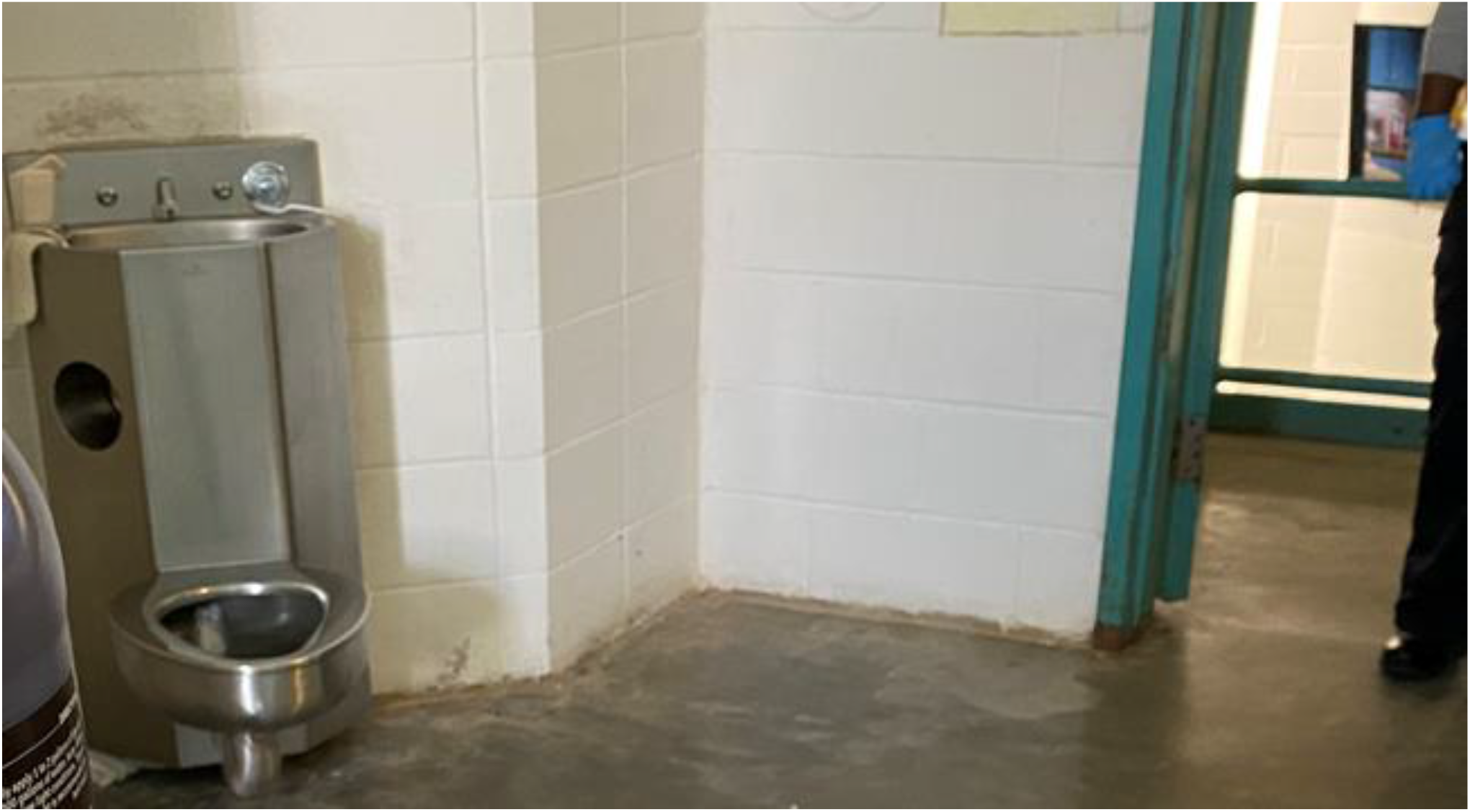
Combined sink-and-toilet. Source: Photograph of author (23)

Despite the information that was gathered from our focus groups, the study had limitations. We conducted this study in just one city, Atlanta, Georgia, although subjects had been incarcerated at many institutions across the state, and also an out-of-state jail. Nonetheless, one study location limits the generalizability of our findings as experiences could vary between counties and states. A second limitation was the short recruitment period of one month because of the urgency of addressing COVID-19. Either a longer recruitment period, more focus groups, or both would have led to a larger sample size. The number of our study subjects was too small to perform deeper analyses such as identify themes by sex, age, and other key demographic features. A more diverse array of experiences and identities would have increased the generalizability of the study as it would apply to a wider population.

Other limitations included recruiting persons formerly detained and incarcerated, rather than currently in a jail. Because of the lengthy process in obtaining IRB approval for involving currently incarcerated persons in research, this decision was made for expediency, but opinions of a person during and after detainment could conceivably change. Also, persons willing to speak with an investigator team may have had less diverse opinions than those borne by a complete cross-section of persons released, thus introducing bias. Lastly, because of the ongoing COVID-19 epidemic, participants wore masks and the interview room doors were left open. This introduced ambient background noise, leading to challenges with acoustics; difficulties hearing one another created problems with sustaining conversations.

This study demonstrates the dividends gained when including justice-involved individuals in the decision-making process regarding interventions for this population. Their input can address factors that could improve the mitigation strategy acceptability. Regarding WBS and self-collection of nasal specimens, their input taught us lessons such as the need to explain clearly what wastewater represents. Among the most compelling findings was the potentially self-empowering role of nasal self-collection. While incarcerated persons seldom have a chance to contribute to decision-making in matters of correctional health, we aspired to address this power imbalance with this present research project.

## Conclusion

We sensed that subjects in this qualitative study found a strategy of pairing wastewater surveillance for SARS-CoV-2 with self-collection of viral specimens acceptable. We thus proceeded with a project based on nasal-self collection and wastewater monitoring for COVID-19 at Fulton County Jail soon after the conclusion of the study, in October 2021.

## Supporting information

SRQR checklist

## Data Availability

Yes - all data are fully available without restriction

## Acknowledgments

We would like to thank the staff members of the center that hosted and facilitated a space for the focus groups, and the subjects for their participation.

## FUNDING

This project was funded by the Bill and Melinda Gates Foundation (INV035562)

## Notes

### Competing Interest Statement

Ms. Gonzalez-Montalvo Mr. Dickson Ms. Saber Ms Boehm and Dr. Phillips declare no competing interests. Dr. Akiyama reports that in the past 5 years he has received grant/project funding through his university from Gilead Sciences and Abbvie. Dr. Spaulding in the past 5 years has received grant/project funding through her university from:
Gilead Sciences
Viral Solutions
Cellex
The National Institutes of Health
The California Department of Corrections Rehabilitation and Health through Walls
She has received direct funding from the National Sheriffs Association and Harris County (Texas). She has received test kits from bioLytical Laboratories through her university for a research project. She has formerly been employed in a part-time capacity by Wellpath and Naphcare.

### Author Declarations

The Emory University IRB approved the study protocol Study # 003291. Participants provided written consent.

