## Supplementary material for "Opinions of Former Jail Residents about Self-collection of SARS-CoV-2 Specimens, Paired with Wastewater Surveillance: A Qualitative Study Rapidly Examining Acceptability of COVID-19 Mitigation Measures": SRQR checklist

### Reporting checklist for qualitative study.

Based on the SRQR guidelines.

#### Instructions to authors

Complete this checklist by entering the page numbers from your manuscript where readers will find each of the items listed below.

Your article may not currently address all the items on the checklist. Please modify your text to include the missing information. If you are certain that an item does not apply, please write "n/a" and provide a short explanation.

Upload your completed checklist as an extra file when you submit to a journal.

In your methods section, say that you used the SRQRreporting guidelines, and cite them as:

O'Brien BC, Harris IB, Beckman TJ, Reed DA, Cook DA. Standards for reporting qualitative research: a synthesis of recommendations. Acad Med. 2014;89(9):1245-1251.

| | | Reporting Item | Page Number |
|---|---|---|---|
| **Title** | | | |
| | #1 | Concise description of the nature and topic of the study identifying the study as qualitative or indicating the approach (e.g. ethnography, grounded theory) or data collection methods (e.g. interview, focus group) is recommended | 1 |
| **Abstract** | | | |
| | #2 | Summary of the key elements of the study using the abstract format of the intended publication; typically includes background, purpose, methods, results and conclusions | 1 |
| **Introduction** | | | |
| Problem formulation | #3 | Description and signifcance of the problem / phenomenon studied: review of relevant theory and empirical work; problem statement | 2 |

| | | | |
|---|---|---|---|
| Purpose or research question | #4 | Purpose of the study and specific objectives or questions | 2 |
| **Methods** | | | |
| Qualitative approach and research paradigm | #5 | Qualitative approach (e.g. ethnography, grounded theory, case study, phenomenolgy, narrative research) and guiding theory if appropriate; identifying the research paradigm (e.g. postpositivist, constructivist / interpretivist) is also recommended; rationale. The rationale should briefly discuss the justification for choosing that theory, approach, method or technique rather than other options available; the assumptions and limitations implicit in those choices and how those choices influence study conclusions and transferability. As appropriate the rationale for several items might be discussed together. | 3-4 |
| Researcher characteristics and reflexivity | #6 | Researchers' characteristics that may influence the research, including personal attributes, qualifications / experience, relationship with participants, assumptions and / or presuppositions; potential or actual interaction between researchers' characteristics and the research questions, approach, methods, results and / or transferability | 3 |
| Context | #7 | Setting / site and salient contextual factors; rationale | 3 |
| Sampling strategy | #8 | How and why research participants, documents, or events were selected; criteria for deciding when no further sampling was necessary (e.g. sampling saturation); rationale | 4-5 |
| Ethical issues pertaining to human subjects | #9 | Documentation of approval by an appropriate ethics review board and participant consent, or explanation for lack thereof; other confidentiality and data security issues | 4 |
| Data collection methods | #10 | Types of data collected; details of data collection procedures including (as appropriate) start and stop dates of data collection and analysis, iterative process, triangulation of sources / methods, and modification of procedures in response to evolving study findings; rationale | 4 |

| | | | |
|---|---|---|---|
| Data collection instruments and technologies | #11 | Description of instruments (e.g. interview guides, questionnaires) and devices (e.g. audio recorders) used for data collection; if / how the instruments(s) changed over the course of the study | 4-5 |
| Units of study | #12 | Number and relevant characteristics of participants, documents, or events included in the study; level of participation (could be reported in results) | 5-6 |
| Data processing | #13 | Methods for processing data prior to and during analysis, including transcription, data entry, data management and security, verification of data integrity, data coding, and anonymisation / deidentification of excerpts | 4-5 |
| Data analysis | #14 | Process by which inferences, themes, etc. were identified and developed, including the researchers involved in data analysis; usually references a specific paradigm or approach; rationale | 5 |
| Techniques to enhance trustworthiness | #15 | Techniques to enhance trustworthiness and credibility of data analysis (e.g. member checking, audit trail, triangulation); rationale | 5 |
| **Results/findings** | | | |
| Syntheses and interpretation | #16 | Main findings (e.g. interpretations, inferences, and themes); might include development of a theory or model, or integration with prior research or theory | 5-8 |
| Links to empirical data | #17 | Evidence (e.g. quotes, field notes, text excerpts, photographs) to substantiate analytic findings | 5-8 |
| **Discussion** | | | |
| Intergration with prior work, implications, transferability and contribution(s) to the field | #18 | Short summary of main findings; explanation of how findings and conclusions connect to, support, elaborate on, or challenge conclusions of earlier scholarship; discussion of scope of application / generalizability; identification of unique contributions(s) to scholarship in a discipline or field | 8-10 |
| Limitations | #19 | Trustworthiness and limitations of findings | 9-10 |

**Other**

| | | | |
|---|---|---|---|
| Conflicts of interest | #20 | Potential sources of influence of perceived influence on study conduct and conclusions; how these were managed | - |
| Funding | #21 | Sources of funding and other support; role of funders in data collection, interpretation and reporting | - |

The SRQR checklist is distributed with permission of Wolters Kluwer © 2014 by the Association of American Medical Colleges. This checklist was completed on 14. June 2019 using https://www.goodreports.org/, a tool made by the EQUATOR Network in collaboration with Penelope.ai
